# Effectiveness of psychosocial interventions for hypertensive disorders in pregnancy: A systematic review and meta-analysis

**DOI:** 10.1101/2022.01.13.22269011

**Authors:** Daniel A. Nnate, Kobi V. Ajayi, Md Mahbub Hossain, Paul Guerby

## Abstract

**Objective:** Studies on psychosocial interventions for perinatal mental health and wellbeing are mostly limited to the postpartum period. However, the physiological changes associated with hypertensive disorders in pregnancy predisposes women to severe psychological distress and adverse birth outcomes. This review investigated the effectiveness of psychosocial interventions for hypertensive disorders during pregnancy.

**Methods:** Cochrane CENTRAL, Embase, MEDLINE, MIDIRS, CINAHL, PsycINFO, PsycArticles, and Web of Science were searched up to 22nd August 2021. Effect sizes on relevant health outcomes were pooled in a meta-analysis using STATA software.

**Results:** Eight randomised trials involving 460 participants met the inclusion criteria. Included studies adopted several interventions ranging from music, exercise, cognitive behavioural therapy (CBT), spiritual care education and psychoeducation. The pooled effect showed a significant reduction in anxiety (d= −0.35 [−0.58, −0.11], p=0.004) and depression (d= −0.37 [−0.57, −0.17], p=0.0003). Spiritual care education significantly reduced postpartum stress disorder (d= −62.00 [−93.10, −30.90], p= 0.0001). However, CBT showed no effect on gestational stress (d= −2.20 [-4.89, 0.48], p= 0.11).

**Conclusion:** This study provides satisfactory evidence that psychosocial interventions may likely reduce anxiety and depression associated with hypertensive disorders in pregnancy. However, the evidence is very uncertain about its effect on neonatal outcomes.

**Summary of findings:** 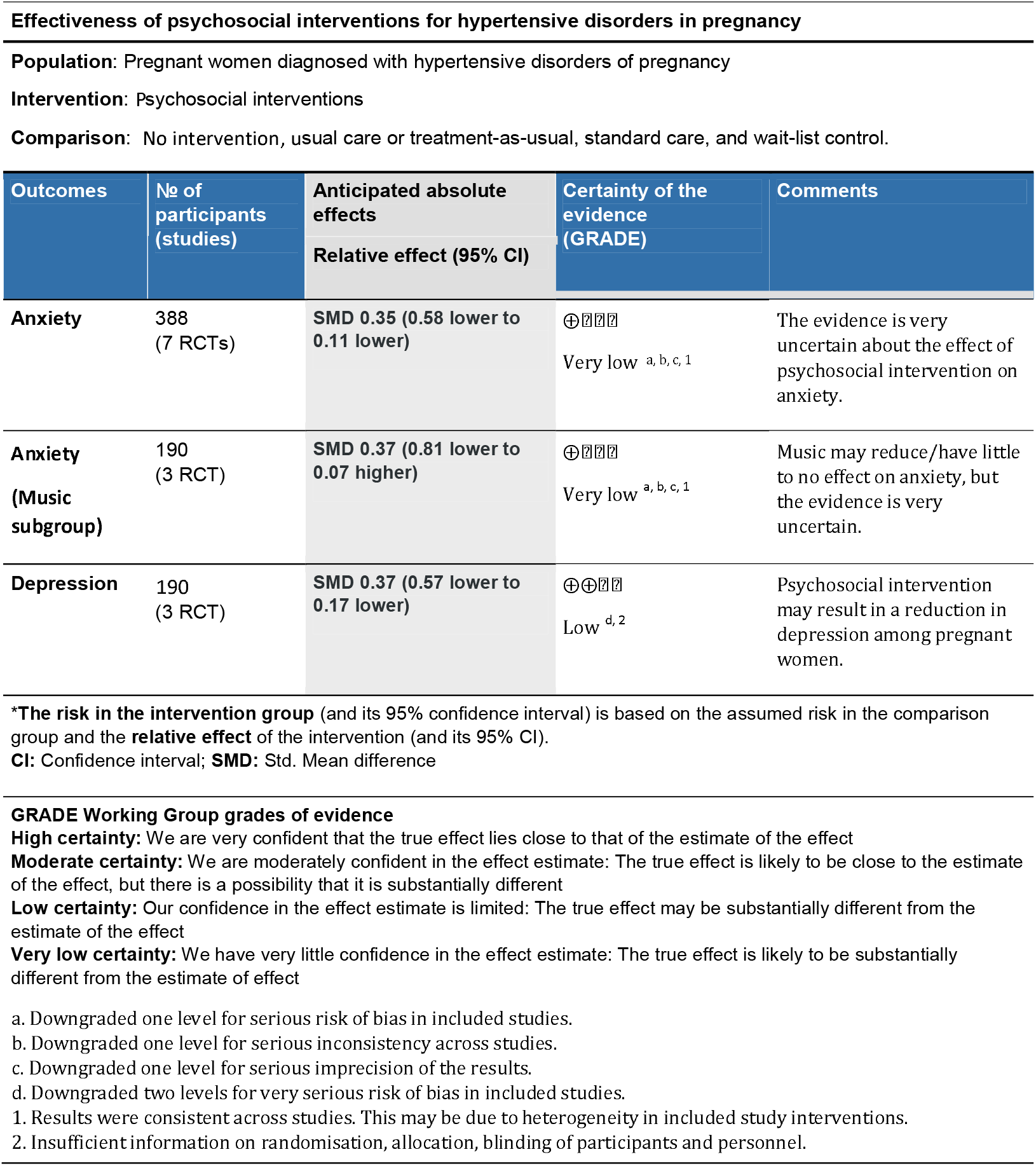

## 1. Introduction

Globally, hypertensive disorders of pregnancy (HDP) are the leading causes of maternal and perinatal morbidity and mortality [1–3]. Hypertensive disorders constitute several entities, including chronic hypertension, gestational hypertension, preeclampsia, and chronic hypertension with superimposed preeclampsia [4]. Although hypertensive disorders are directly associated with about 5-10% of adverse pregnancy outcomes [5], preeclampsia and eclampsia contribute to the greatest burden of deaths [6, 7]. Risk factors across the disorders that predispose women include maternal age (over 40 years), primigravida pregnancy, family history of hypertension and pre-eclampsia, body mass index (BMI) above 30kg/m^2^, hormone imbalance, multiple-birth pregnancy, pregnancy interval greater than 10 years; assisted reproductive technologies and pre-existing medical conditions like diabetes, renal disease, and autoimmune disorders such as systemic lupus erythematosus [4, 8, 9]. These chronic diseases may also be associated with increased anxiety, depression, and poor quality of life in pregnant women.

Research indicates that HDP is associated with increased maternal mental health burdens such as depression and anxiety [2, 10–12]. Although depression and anxiety are common among pregnant women, leading to postpartum depression and/or anxiety and other mental health problems, combining these conditions with HDP is especially detrimental to maternal and infant health. A meta-analysis of 61.2 million pregnancies suggested that depression and anxiety were clinically associated with HDP [11]. Specifically, this study reported that women who met the clinical cut-off score for depression and/or anxiety symptoms had a 39% increased relative risk of HDP diagnosis than those without these conditions [11].

There are several psychosocial (e.g., yoga, meditation, physical activities) and psychological interventions (e.g., cognitive behavioural therapy [CBT]), psychoeducation, and psychotherapy) to prevent or to treat perinatal depression and anxiety [13–16]. Evidence suggests that these therapeutic interventions can prevent or reduce depression and anxiety symptoms among pregnant women and postpartum women up to one year after childbirth.

However, a summary of the literature on the effectiveness of psychosocial interventions among women with hypertensive disorders is lacking. Nonetheless, there is evidence that physiological and psychological changes associated with pregnancy predispose women to anxiety and depression which causes fear of childbirth and poor birth outcomes and postnatal depression in the early postpartum period [17–19].

Although clinical evidence supports the linkage between psychological distress and HDP [20, 21]; the need for stress management interventions for pregnant women has also been emphasised in international guidelines [1, 2]. Given the evidence about psychosocial interventions among women with HDP, summarizing and synthesizing the literature is significant to inform practice on mitigating the negative consequences of mental disorders on maternal and foetal health. In addition, there is a need to ascertain which psychosocial interventions could best ameliorate psychological distress, depression, and anxiety among women diagnosed with HDP. Therefore, the purpose of this study is to summarize the existing literature reporting the effectiveness of psychosocial interventions for improving mental health outcomes in pregnancies complicated by hypertensive disorders.

## 2. Methods

The protocol for this systematic review was registered with PROSPERO (registration number: CRD42020208981). The review is reported following the Preferred Reporting Items for Systematic Review and Meta-Analysis (PRISMA 2020) guidelines [22]. Ethics approval was not required for this systematic review and meta-analysis.

### Eligibility Criteria

#### Criteria for studies

Studies on pregnant women diagnosed with a hypertensive disorders were defined according to the criteria of the American College of Obstetricians and Gynaecologists (ACOG) and the International Society for the Study of Hypertension in Pregnancy (ISSHP) for gestational hypertension: elevated systolic blood pressure (SBP) ≥140 mm Hg or diastolic blood pressure (DBP) ≥ 90 mm Hg after 20 weeks of gestation in a previously non-hypertensive woman; preeclampsia: an elevated SBP ≥140 mm Hg or DBP ≥90 mm Hg in a previously non-hypertensive woman accompanied by proteinuria (excretion of ≥ 0.3 g protein every 24 hour); chronic hypertension, defined as SBP ≥140 mm Hg or DBP ≥90 mm Hg before 20 weeks of gestation; HELLP (haemolysis, elevated liver enzymes, low platelets) syndrome defined by thrombocyte count <100 × 10^9^/L, and/or ASAT and ALAT ≥30 U/L), and superimposed preeclampsia, an increase in BP and a new onset of proteinuria with previous hypertension before 20 weeks of gestation accompanied by HELLP syndrome [23–25].

### Intervention

All studies that evaluated the effect of psychosocial interventions, including psychotherapy (e.g., meditation), psychological interventions (e.g., cognitive behaviour therapy (CBT), mindfulness-based intervention), exercise and other psychosocial interventions (e.g., breathing exercise, physical activity, and yoga) in women with hypertensive disorders during the perinatal periods were considered.

### Comparator

Intervention recipients were compared to controls who received usual care or treatment-as-usual, standard care, wait-list control, placebo or other active controls or interventions.

### Outcomes

Primary outcomes of interest were focused on maternal mental health disorders such as anxiety, depression, and psychological distress. Any reported undesirable effects in the neonate such as gestational stress and low birth weight were also considered. Selected study outcomes were further restricted to those collected from validated questionnaires.

### Study type

Randomised controlled studies comparing one or more interventions with a control group with or without blinding.

### Search strategy

We initially identified index terms and keywords based on the participants, intervention, comparator, outcome, and study design (PICOS) framework. The search string was then refined for greater sensitivity by excluding keywords for comparison and outcome (see Supplementary Table S1). Electronic searches were drawn up from Cochrane Central Register of Controlled Trials (CENTRAL) Pregnancy and Childbirth Group, Embase (OVID), MEDLINE (Ovid), MIDIRS (Ovid), PsycINFO (Ovid), CINAHL (EBSCOhost), PsycArticles (ProQuest) and Web of Science up to 22nd August 2021. Hand searching was also carried out on Google Scholar and journals related to obstetrics and gynaecology not indexed in bibliographic databases. Relevant studies were also extracted from reference lists of identified studies. No restrictions on language, date and age of participants were applied.

### Study selection and data extraction

Identified records were exported to Covidence, online software for screening and data extraction [26]. After removing duplicate studies, titles and abstracts were first screened independently by DAN and KVA to determine the suitability of the studies using keywords developed from the population, intervention, comparator, and outcome (PICOS) framework. All identified studies from the searched databases were checked by DAN and KVA and assessed against the inclusion and exclusion criteria. Disagreements were resolved through discussion with MMH and PG who performed an additional independent evaluation. Data extracted from eligible studies encompassed study characteristics (first author, year of publication, country, and sample size); intervention (the type of intervention and control, duration of intervention in weeks); patient characteristics (mean age and current treatment); study outcomes and treatment duration (see Table 1).

**TABLE 1.**
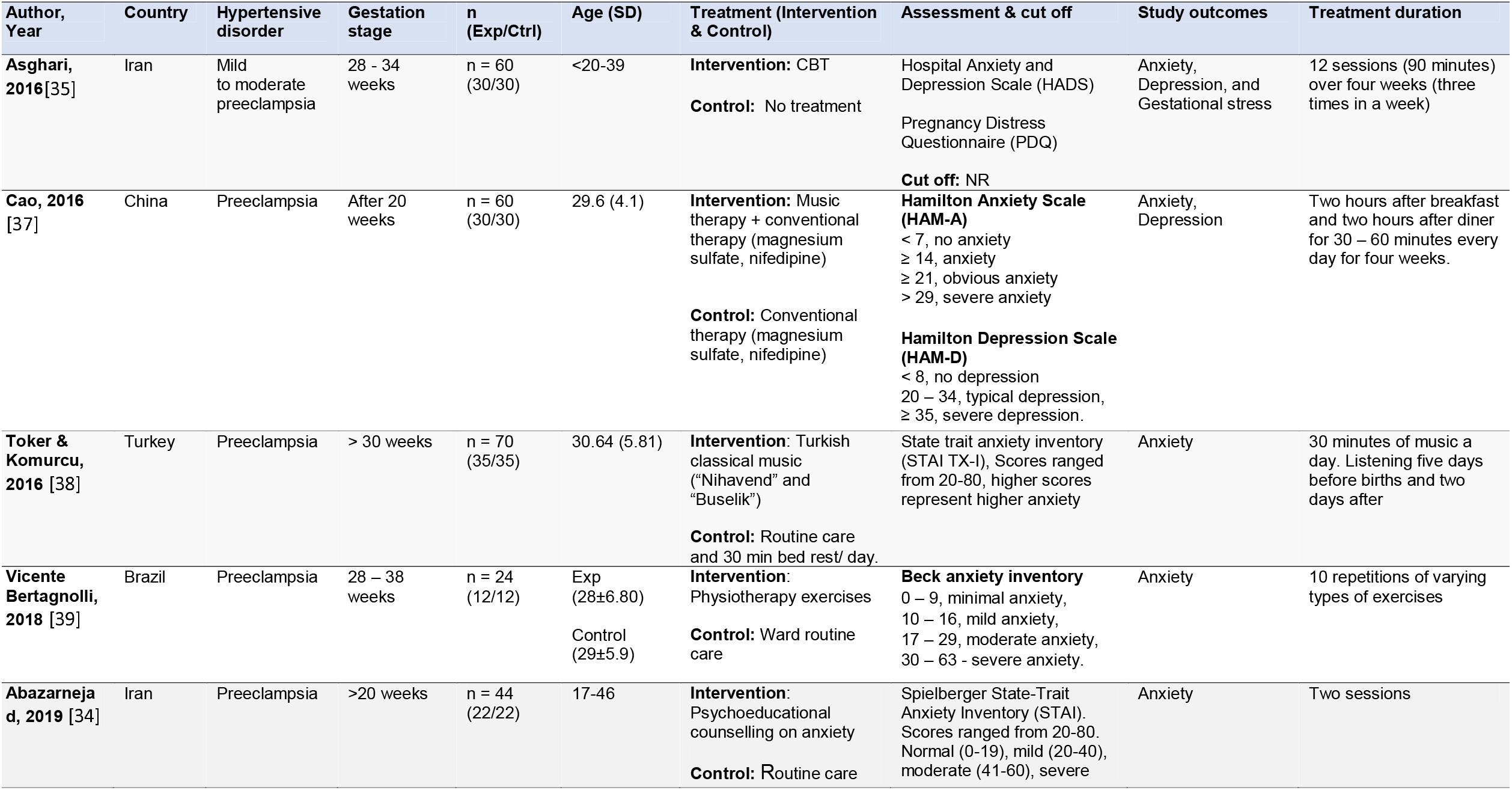
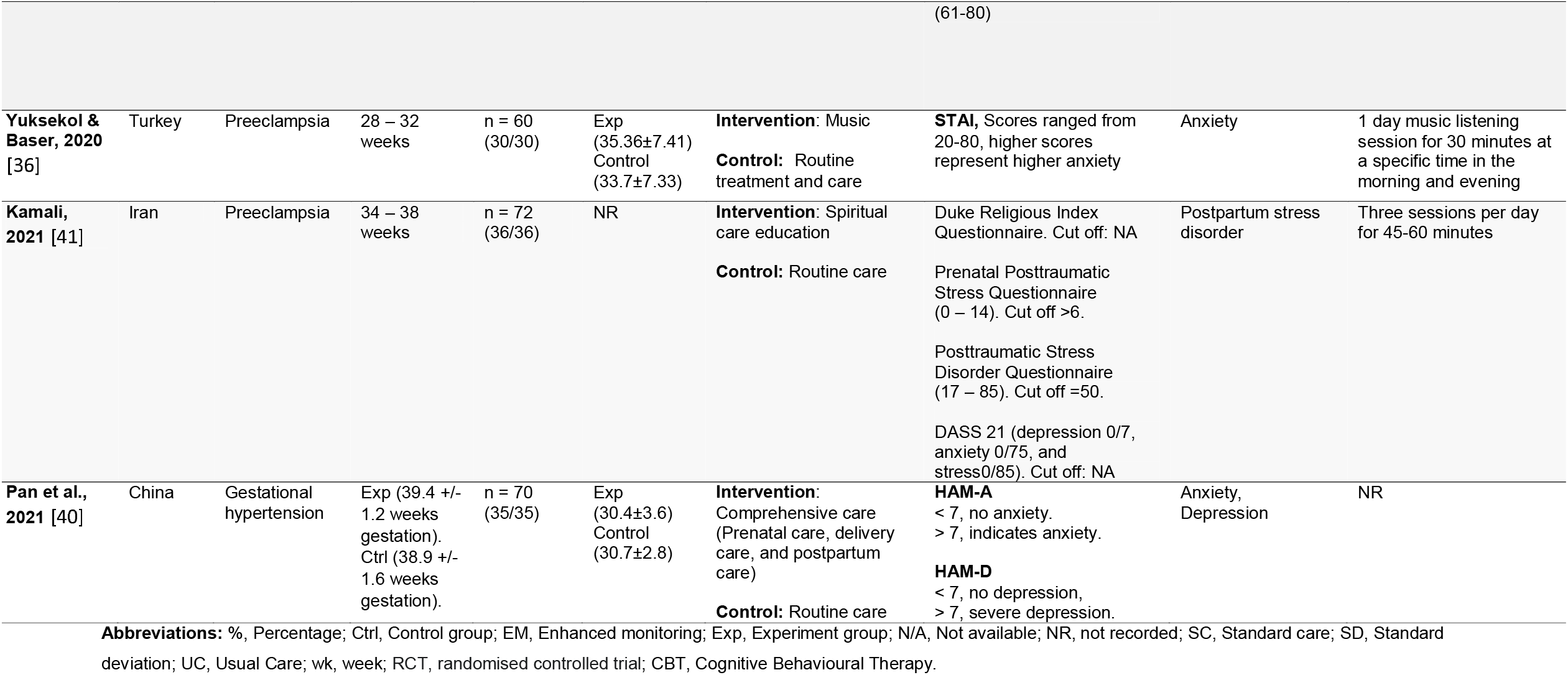
Characteristics of included studies

### Risk of bias and quality assessment

A modified version of the Effective Practice and Organisation of Care (EPOC) risk of bias (RoB) tool for randomised trials was used for quality assessment of included studies [27, 28]. Included studies were assessed for bias in the following domain: selection bias (difference between baseline characteristics, poor random sequence generation and allocation concealment), detection bias (different outcome assessment), performance bias (blinding of participants, study personnel and outcome assessment), attrition bias (incomplete outcome data) and reporting bias (selective reporting of outcomes and publication bias or spin). RoB in each domain and within each study was rated as low, unclear, or high (see Supplementary Table S3).

### Data analysis

Study outcomes were pooled in a meta-analysis using STATA software (Version 16). Where included studies adopted different scales for measuring outcomes of interest, standardise mean differences of reported outcomes were generated before pooling in a meta-analysis. The effect size was expressed as Cohen’s d [95% confidence intervals]. Pooled results were represented graphically as forest plots and statistical significance was defined as p-value ≤ 0.05. Subgroup analysis was carried out to determine the effect of selected interventions on various outcomes. I-squared (I^2^) values less than 50% implies a low level of heterogeneity among the intervention effects, while an I^2^ value greater than 50% signified a large degree of heterogeneity [[29, 30]. Where I^2^ is greater than 50%, the DerSimonian-Laird random-effects model was recommended [29]. Publication bias in included studies was investigated using Egger’s tests for the symmetry [31]. A p-value less than 0.05 implies the presence of publication bias. The GRADE approach was used to assess the certainty of clinically relevant outcomes, and a summary of findings table was further created using the GRADEpro Guideline Development Tool (GDT) [32, 33] (See summary of findings table).

## 3. Results

### Study selection

The approach used for selecting included studies was summarised in a PRISMA flow diagram to ensure transparency in the screening and study selection process (see Figure 1). The search strategy yielded 492 citations with 45 hits in Cochrane CENTRAL, 48 in Embase; 21 in MEDLINE; 34 in MIDIRS; 76 in CINHAL; 44 in PsycINFO; 56 in PsycArticles and 167 from Web of Science. After the removal of duplicates and irrelevant records, a total of 15 full-text articles were retained and screened for eligibility. Seven studies were further excluded with reasons for the exclusion provided. In total, 8 RCTs met the inclusion criteria.

**Figure 1.**
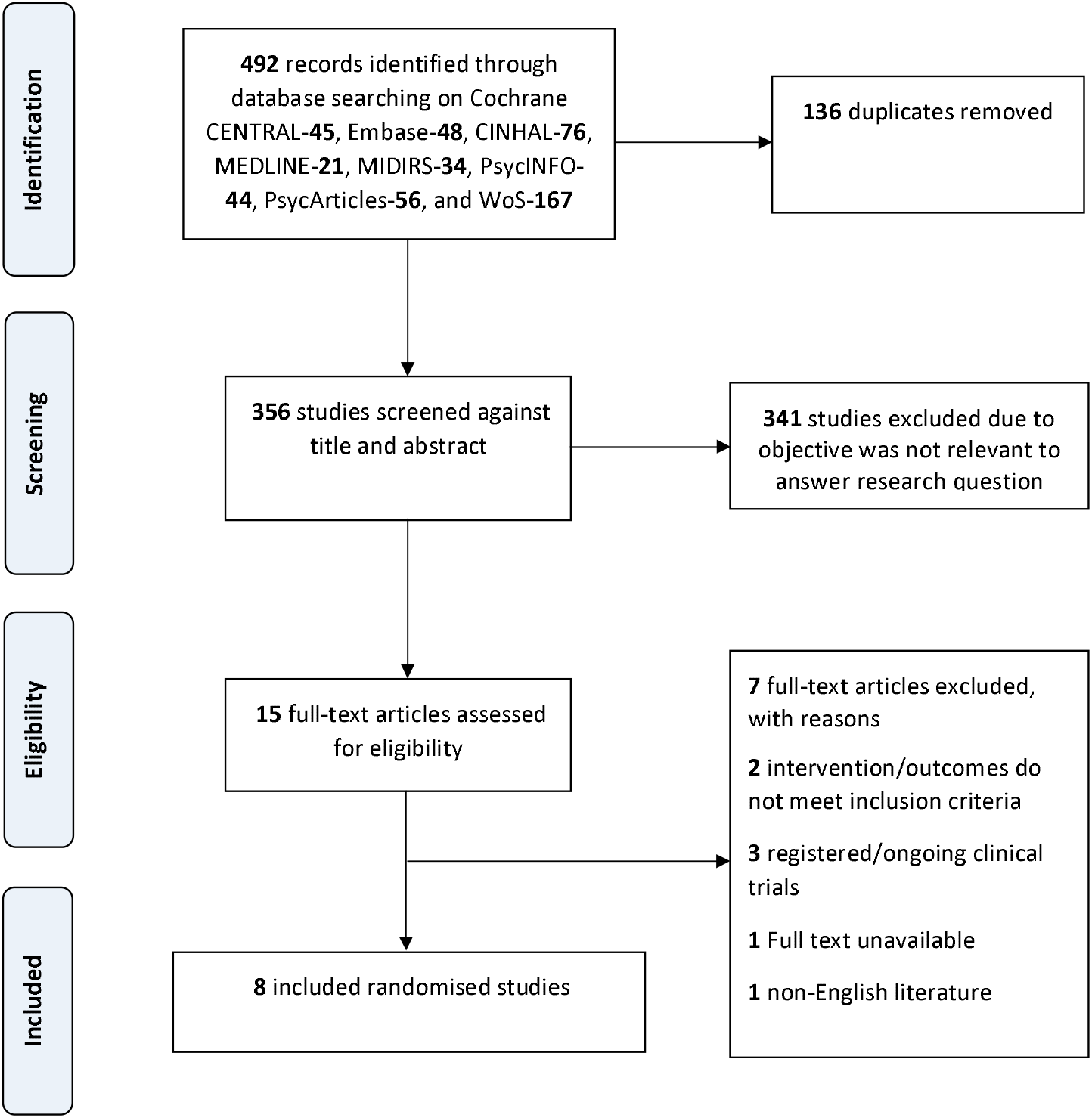
PRISMA flow diagram of the study selection procedure.

### Characteristics of included studies

The psychosocial interventions used in the included studies ranges from psychoeducational counselling [34], cognitive behavioural therapy (CBT) [35], music therapy [36, 37], Turkish classical music [38], physiotherapy exercises [39], comprehensive care [40], and spiritual care education [41]. (see Table 1). The study by Vicente Bertagnolli et al. [39] was carried out on participants in Brazil. Three studies [34, 35, 41] were conducted in Iran, two [36, 38] in Turkey and another two studies [37, 40] were undertaken in China. The average age of the participants was 31.5 years; range,17-46 years. The sample sizes of the included studies ranged between 24 and 72. Of the 460 pregnant women, 390 of them had preeclampsia [34–39, 41] and 70 had gestational hypertension [40]. The average gestational age of participants was 29.7 weeks; range, 20-39 weeks. The questionnaires utilised for measuring anxiety, depression, psychological distress and any undesirable outcome on neonate is presented in Table 1.

### Risk of bias and quality of included studies

The risk of bias in randomised trials was based on the modified 9-item EPOC tool which allowed for an enhanced assessment of study participants and outcomes [28]. (See Supplementary Table S2). The EPOC RoB tool was divided into two sections. In the first section, participants and personnel were assessed for selection and performance bias. Outcomes of study that merit inclusion after stage one was further assessed for selective outcome reporting, similar baseline outcome measurements and incomplete outcome data as recommended by Nnate et al. [28]. Studies that record a ‘high risk’ in the first section were of poor quality and pose further RoB in the outcome assessment. Studies were graded as low, unclear, or high risk. Overall, the current review was rated as ‘moderate quality’. A summary of RoB within included studies was presented in Table 2.

**Table 2.**
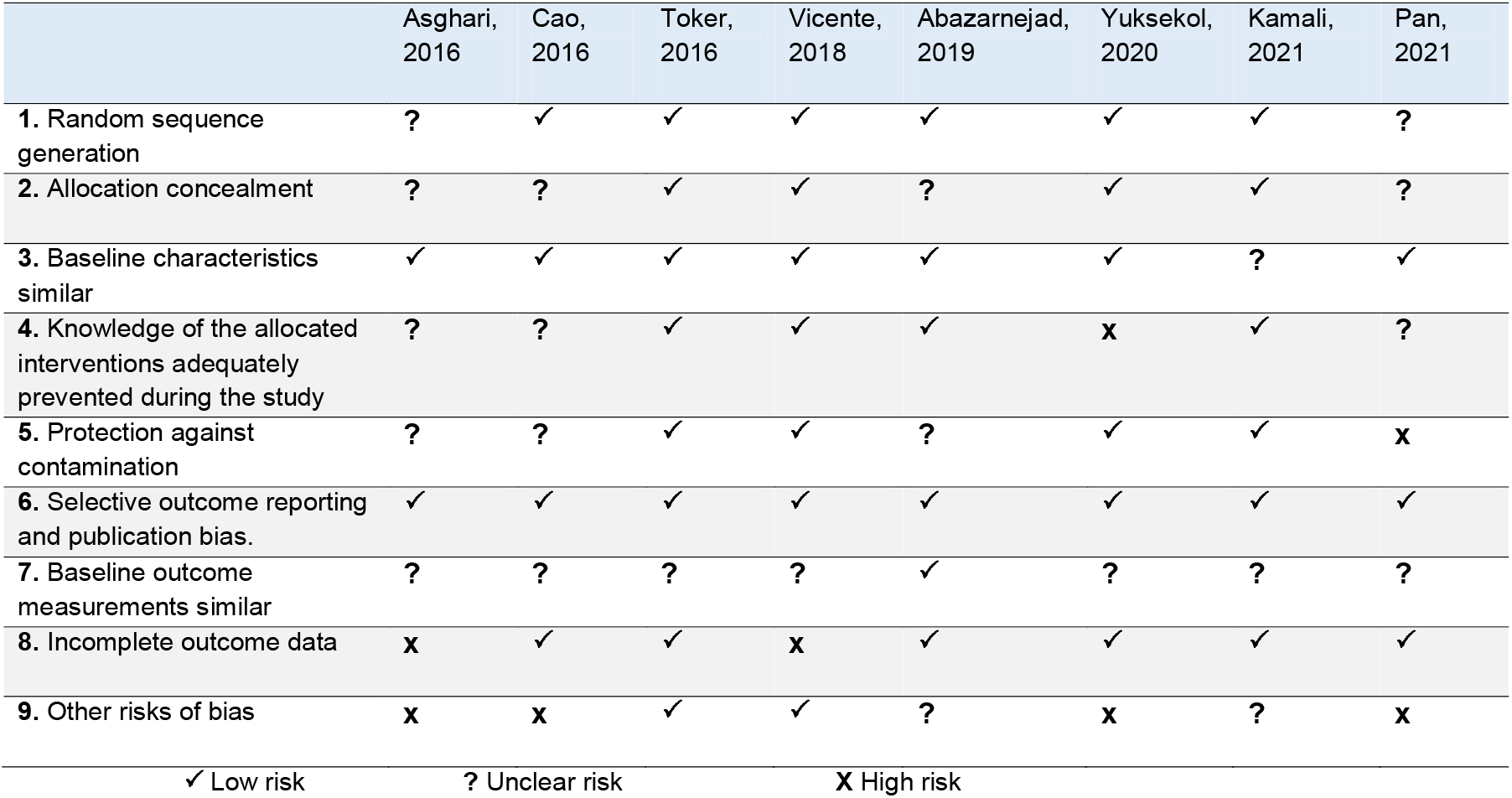
Risk of bias assessment within included studies

|  | Asghari,<br>2016 | Cao,<br>2016 | Toker,<br>2016 | Vicente,<br>2018 | Abazarnejad,<br>2019 | Yuksekol,<br>2020 | Kamali,<br>2021 | Pan,<br>2021 |
| --- | --- | --- | --- | --- | --- | --- | --- | --- |
| 1. Random sequence generation | ? | ✓ | ✓ | ✓ | ✓ | ✓ | ✓ | ? |
| 2. Allocation concealment | ? | ? | ✓ | ✓ | ? | ✓ | ✓ | ? |
| 3. Baseline characteristics similar | ✓ | ✓ | ✓ | ✓ | ✓ | ✓ | ? | ✓ |
| 4. Knowledge of the allocated interventions adequately prevented during the study | ? | ? | ✓ | ✓ | ✓ | x | ✓ | ? |
| 5. Protection against contamination | ? | ? | ✓ | ✓ | ? | ✓ | ✓ | x |
| 6. Selective outcome reporting and publication bias. | ✓ | ✓ | ✓ | ✓ | ✓ | ✓ | ✓ | ✓ |
| 7. Baseline outcome measurements similar | ? | ? | ? | ? | ✓ | ? | ? | ? |
| 8. Incomplete outcome data | x | ✓ | ✓ | x | ✓ | ✓ | ✓ | ✓ |
| 9. Other risks of bias | x | x | ✓ | ✓ | ? | x | ? | x |
| ✓ Low risk<br>? Unclear risk<br>x High risk |  |  |  |  |  |  |  |  |

### Study outcomes

Included study interventions were administered at different frequencies. In studies that utilized music, the duration was between 30-60 minutes. However, while Cao et al. [37] and Toker et al. [38] administered music for longer days, Yuksekol et al. [36] was only for a day. Asghari et al. [35] delivered 12 sessions of a 90-minutes CBT over 4 weeks period. In Kamali et al. [41], participants received 3 sessions of spiritual care education for 45-60 minutes but there was no report on any repetitions. Additionally, participants received 10 repetitions of varying types of exercises in Vicente Bertagnolli et al. [39] but there was no report on its duration. While Abazarnejad et al. [34] delivered 2 psychoeducational counselling sessions, Pan et al. [40] on the other hand did not report any information on treatment duration.

Included studies that evaluated anxiety [34–40] and depression [35, 37, 40] adopted different interventions. The level of heterogeneity was moderate for anxiety (I^2^ = 62.44%) and homogenous for depression (I^2^ = 0%). Therefore, pooled results were reported as standardised means using the DerSimonian-Laird random-effects model where I^2^ >50 and inverse-variance fixed-effect where I^2^ =0. (see Figures 2 and 3). There was significant reduction in anxiety (d= −0.35 [−0.58, −0.11], p=0.004) in pregnant women with HDP. Egger’s test (z = 0.68, p = 0.4937) revealed no significant publication bias. Psychosocial intervention also reduces depression (d= −0.37 [−0.57, −0.17], p=0.0003) in pregnant women with HDP. Egger’s test (z = 0.58, p = 0.5604) revealed no significant publication bias.

**Figure 2.**
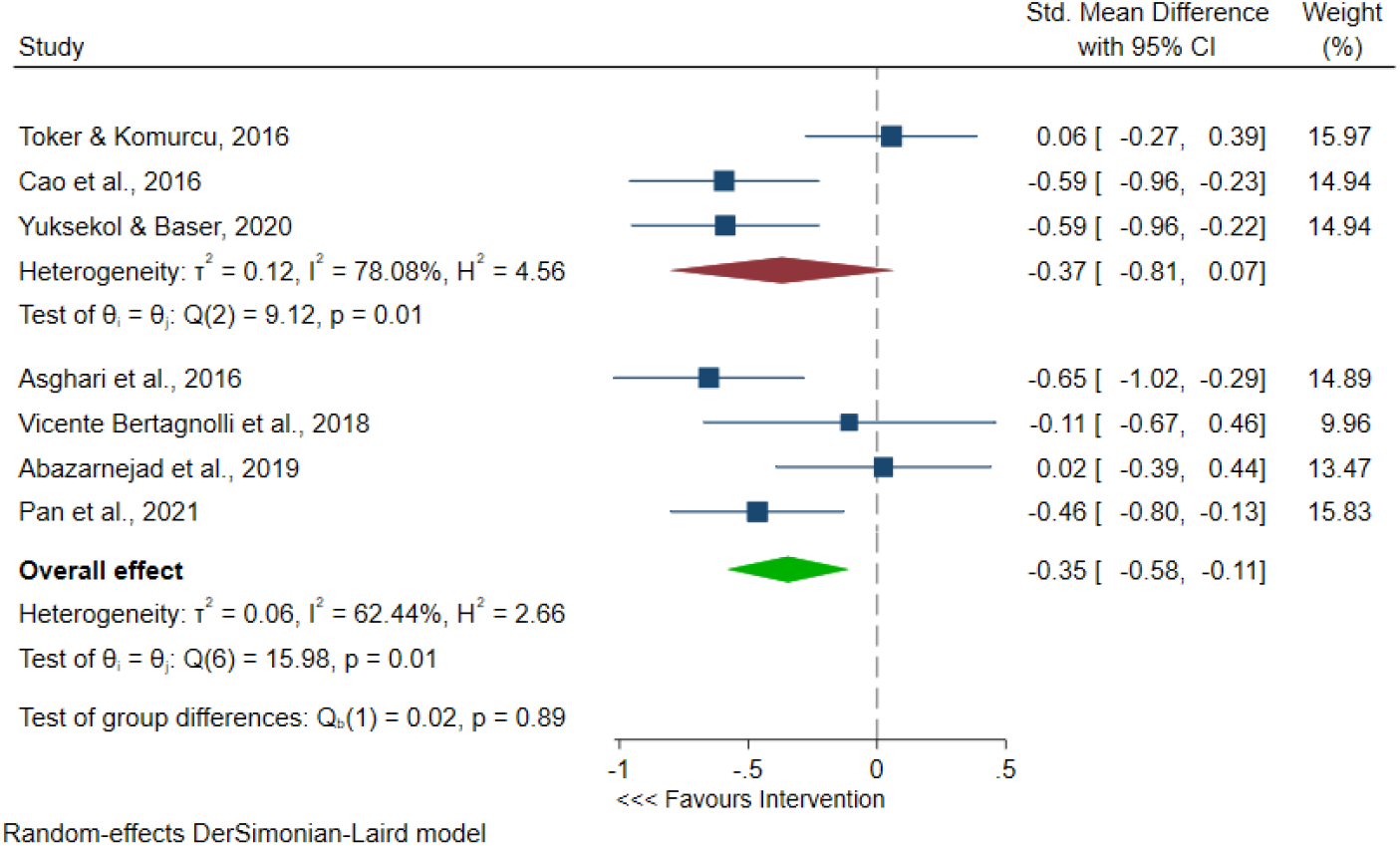
Forest plot of pooled effect estimate for anxiety

**Figure 3.**
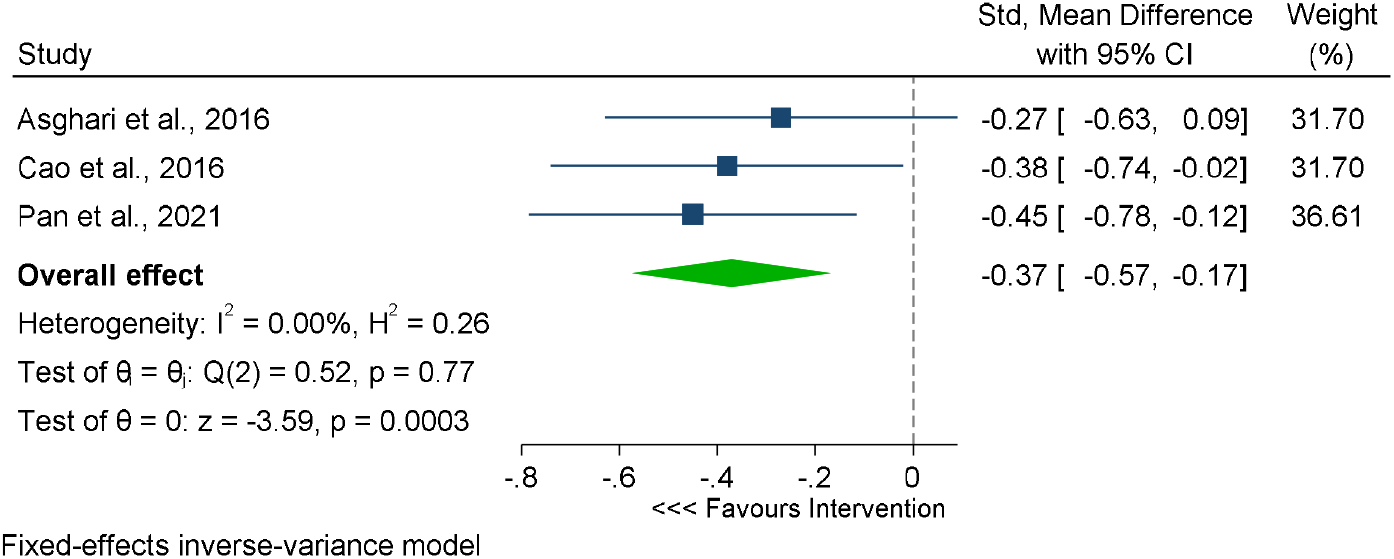
Forest plot of pooled effect estimate for depression

Kamali et al.[41] also reported a significant reduction in postpartum stress disorder with spiritual care education (d= −62.00 [−93.10, −30.90], p=0.0001). In the study by Asghari et al. [35], CBT showed little effect on gestational stress, but the evidence presented is very uncertain (d= −2.20 [-4.89, 0.48], p=0.11). Forest plots for postpartum stress disorder and gestational stress were illustrated separately in Figure 4.

**Figure 4.**
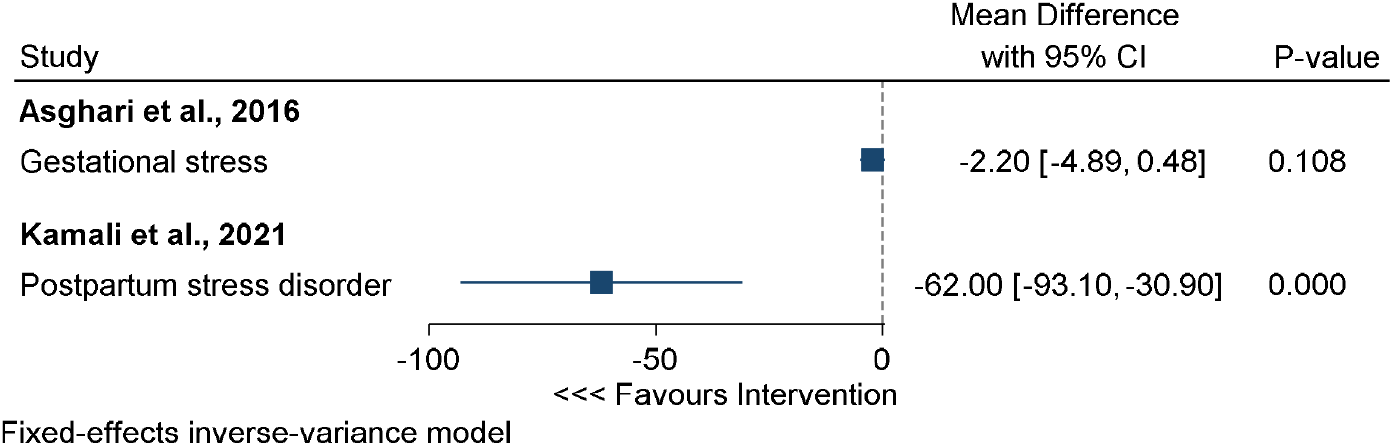
Forest of effect estimate for gestational stress and postpartum stress disorder.

## 4. Discussion

The results of this systematic review and meta-analysis show that adverse perinatal mental health conditions that are associated with HDP could be decreased using various psychosocial interventions. Low to medium effect sizes for multiple mental health outcomes were significant, which suggest the efficacy of psychosocial interventions within the context of these interventions. However, the evidence also informs high heterogeneity within a low number of studies, which may affect the generalizability of the findings.

The level of heterogeneity observed for anxiety could be related to several factors ranging from the difference in gestational age, age of participants, type of study intervention, treatment duration and assessment tools with different cut-offs. A subgroup analysis for music intervention revealed a significant decrease in anxiety in participants with preeclampsia (see Figure 2). Turkish classical music [38] had no effect on anxiety contrary to the results of music therapy [36, 37]. Likewise, psychoeducational counselling [34] resulted in little to no reduction in anxiety. Although the majority of participants in the included studies were diagnosed with preeclampsia, [34–39, 41] there was also no relationship between the prognosis and the achieved effect on participants. Similarly with depression, there was insufficient evidence to make any causal relationship regarding the observed homogeneity in study results.

However, the findings of the current study may not support the previous research suggesting that psychoeducation was effective in mitigating anxiety and fear of childbirth in pregnant women [18, 19]. The study by Kamali et al.[41] and Asghari et al.[35] assessed the effect of spiritual care education and CBT on postpartum stress disorder and gestational distress respectively. While there was a significant decrease in postpartum stress disorder, CBT may not reduce gestational distress in neonates. Although research evidence shows that psychosocial interventions significantly improved postnatal depression [42], there was a lack of evidence to suggest that CBT was effective in preventing gestational distress.

A recent study on the effect of CBT on gestational age at birth and birth weight in pregnant women with an anxiety disorder also reported a similar result [43]. While the authors reported a positive effect of CBT on perinatal mental health, there was no significant difference in gestational age or birth weight between the treatment and control groups [43]. This finding is contrary to that of Bleker et al. [44] which showed a positive effect on neurobiological, behavioural and cognitive outcomes in children over 3 years postpartum. Therefore, considering current evidence, it is very uncertain about the effect of psychosocial interventions on offspring.

A striking feature observed in this study was the positive effect of CBT, music therapy and comprehensive care on depression [35, 37, 40]. This may suggest that pregnant women with hypertensive disorders are likely to benefit from these interventions. However, comprehensive care intervention by Pan et al. [40] was defined as a combination of prenatal care, delivery care, and postpartum care, we are uncertain of the effect of delivery care and postpartum care on depression in participants with gestational hypertension. Nevertheless, the group of interventions that made up prenatal care included diet modification, counselling and emotional self-regulation. Another drawback of the study by Pan et al. [40] was that the authors were not specific on what made up prenatal care. Therefore, it is important to bear in mind the possible bias in the study results.

The template for intervention description and replication (TIDieR) guide has been recommended for better reporting of interventions in published papers [45]. This is because the significance of an intervention, and whether research findings may be implemented or replicated in another group of individuals depends on the description of the interventions. The 12-item TIDieR checklist and guide improves the reporting of interventions and makes it easier for researchers to structure reports of their interventions in order for readers to understand. Authors of seven [34–39, 41] of the included studies discussed in detail the nature of their interventions. The study by Pan et al.[40] evaluated the effect of comprehensive care, however, only listed a group of interventions delivered to participants. In general, there was a poor description of treatment duration in some of the included studies as well [34, 39–41]. Therefore, it may not be feasible to replicate such studies or adopt the interventions in clinical settings.

Previous studies have revealed an upward trend in anxiety and depression across trimesters [19, 46]. Nevertheless, our study has failed to show if there were more uptake of these interventions in the third trimester. While the included studies had a higher number of participants diagnosed with preeclampsia, the findings of this review will be of interest to clinical psychologists providing care to women with preeclampsia. Furthermore, the inclusion of only randomised studies for the meta-analysis further strengthens the findings of this review.

Despite these strengths, the current study has several limitations that should be acknowledged for interpreting the findings and advancing future research and practice. This meta-research found a limited number of trial studies with a low number of participants in those trials that may have affected the effect sizes and heterogeneity. More trials with large and diverse samples are needed to truly understand the efficacy and effectiveness of psychosocial interventions for HDP. Another limitation was the use of different measurement instruments and cut-off values for mental health outcomes within the included trials. A standardized and consistent measurement could have informed altered effect sizes from respective studies, which could change the meta-analytic findings as well. Lastly, we evaluated literature from major databases, however, it is possible that unpublished studies with varying effect sizes remained beyond the scope of this study. Evidence synthesis must be as much inclusive as possible. Therefore, future evidence-based reviews should examine additional literature whenever possible.

## Conclusion

The present study was designed to determine the effectiveness of psychosocial interventions for HDP. Our review further suggests that psychological interventions during pregnancy, and in particular music therapy is likely to reduce maternal anxiety in pregnancies complicated with preeclampsia. Despite the relatively limited evidence, this work also offers valuable insights into probable interventions for reducing anxiety and depressive symptoms associated with HDP. Although CBT resulted in a decrease in anxiety and depression during perinatal periods, the issue of gestational stress is an intriguing one which could be usefully explored in further research. Considerably more work will need to be done to determine the effect of psychosocial interventions on neonatal outcomes.

## Data Availability

All data produced are available online at

https://doi.org/10.6084/m9.figshare.17810036.v2

## Data availability statement

The data that support the findings of this study are openly available in Figshare at https://doi.org/10.6084/m9.figshare.17810036.v2, reference number [47].

