## Supplementary Table S1 for "Effectiveness of psychosocial interventions for hypertensive disorders in pregnancy: A systematic review and meta-analysis"

**Appendix S1. Search strategy**

**Embase 1974 to 2021 August 20**

| # | **Searches** | **Results** |
| --- | --- | --- |
| 1 | exp cognitive behavioural therapy/ or cognitive behavioural therapy or CBT.mp. | 33055 |
| 2 | exp psychotherapy/ or psychotherapy.mp. or therap*.mp. | 9293716 |
| 3 | (psychological or behavio*ral or psych*social).mp. | 1363123 |
| 4 | exp mindfulness/ or mindfulness.mp. | 14578 |
| 5 | exp breathing exercise/ or "breathing exercise".mp. or "breathing therapy".mp. | 8412 |
| 6 | exp "acceptance and commitment therapy"/ or (acceptance and commitment therapy).mp. | 360267 |
| 7 | exp meditation/ or meditation.mp. | 11127 |
| 8 | ("dialectical behavio*r therapy" or "metacognitive therapy" or "schema therapy").mp. | 1935 |
| 9 | exp social support/ or "social support".mp. | 111925 |
| 10 | (counsel*ing or "psycho*education").mp. | 228676 |
| 11 | (exercise or "physical activity" or "mind*body" or yoga).mp. | 697223 |
| 12 | 1 or 2 or 3 or 4 or 5 or 6 or 7 or 8 or 9 or 10 or 11 | 11126646 |
| 13 | exp preeclampsia/ or pre*eclampsia.mp. or "maternal hypertension".mp. or "pregnancy*induced hypertension".mp. or "gestational hypertension".mp. | 74704 |
| 14 | ("psychological distress" or anxiety or depression or "mental health" or resilience).mp. | 1195648 |
| 15 | 11 and 12 | 1911 |
| 16 | 12 and 15 | 1146 |
| 17 | limit 14 to human | 1115 |
| 18 | limit 17 to female | 683 |
| 19 | 18 not (meta-analysis or "systematic review" or conference abstract or "conference review" or editorial or commentary or correspondence or letter or note or review or book review or news).mp. | 429 |
| 20 | limit 19 to (clinical trial or randomized controlled trial or controlled clinical trial or multicenter study) [Limit not valid in APA PsycInfo; records were retained] | 48 |

[mp = title, abstract, original title, name of substance word, subject heading word, floating sub-heading word, keyword heading word, organism supplementary concept word, protocol supplementary concept word, rare disease supplementary concept word, unique identifier, synonyms]

**Ovid MEDLINE(R) ALL 1946 to August 20, 2021**

| # | **Searches** | **Results** |
| --- | --- | --- |
| 1 | exp cognitive behavioural therapy/ or cognitive behavioural therapy or CBT.mp. | 14785 |
| 2 | exp psychotherapy/ or psychotherapy.mp. or therap*.mp. | 6574738 |
| 3 | (psychological or behavio*ral or psych*social).mp. | 1036247 |
| 4 | exp mindfulness/ or mindfulness.mp. | 9844 |
| 5 | exp breathing exercise/ or "breathing exercise".mp. or "breathing therapy".mp. | 4111 |
| 6 | exp "acceptance and commitment therapy"/ or (acceptance and commitment therapy).mp. | 306785 |
| 7 | exp meditation/ or meditation.mp. | 6740 |
| 8 | ("dialectical behavio*r therapy" or "metacognitive therapy" or "schema therapy").mp. | 1209 |
| 9 | exp social support/ or "social support".mp. | 97047 |
| 10 | (counsel*ing or "psycho*education").mp. | 132307 |
| 11 | (exercise or "physical activity" or "mind*body" or yoga).mp. | 446779 |
| 12 | 1 or 2 or 3 or 4 or 5 or 6 or 7 or 8 or 9 or 10 or 11 | 7990481 |
| 13 | exp preeclampsia/ or pre*eclampsia.mp. or "maternal hypertension".mp. or "pregnancy*induced hypertension".mp. or "gestational hypertension".mp. | 43906 |
| 14 | ("psychological distress" or anxiety or depression or "mental health" or resilience).mp. | 789551 |
| 15 | 11 and 12 | 465 |
| 16 | 12 and 15 | 254 |
| 17 | limit 14 to human | 220 |
| 18 | limit 17 to female | 217 |
| 19 | 18 not (meta-analysis or "systematic review" or conference abstract or "conference review" or editorial or commentary or correspondence or letter or note or review or book review or news).mp. | 136 |
| 20 | limit 19 to (clinical trial or randomized controlled trial or controlled clinical trial or multicenter study) [Limit not valid in APA PsycInfo; records were retained] | 21 |

[mp = title, abstract, heading word, drug trade name, original title, device manufacturer, drug manufacturer, device trade name, keyword, floating subheading word, candidate term word]

**APA PsycInfo 1806 to August Week 3 2021**

| # | **Searches** | **Results** |
| --- | --- | --- |
| 1 | exp cognitive behavioural therapy/ or cognitive behavioural therapy or CBT.mp. | 17263 |
| 2 | exp psychotherapy/ or psychotherapy.mp. or therap*.mp. | 668844 |
| 3 | (psychological or behavio*ral or psych*social).mp. | 839761 |
| 4 | exp mindfulness/ or mindfulness.mp. | 16716 |
| 5 | exp breathing exercise/ or "breathing exercise".mp. or "breathing therapy".mp. | 135 |
| 6 | exp "acceptance and commitment therapy"/ or (acceptance and commitment therapy).mp. | 79991 |
| 7 | exp meditation/ or meditation.mp. | 9351 |
| 8 | ("dialectical behavio*r therapy" or "metacognitive therapy" or "schema therapy").mp. | 3053 |
| 9 | exp social support/ or "social support".mp. | 85909 |
| 10 | (counsel*ing or "psycho*education").mp. | 116092 |
| 11 | (exercise or "physical activity" or "mind*body" or yoga).mp. | 92864 |
| 12 | 1 or 2 or 3 or 4 or 5 or 6 or 7 or 8 or 9 or 10 or 11 | 1591007 |
| 13 | exp preeclampsia/ or pre*eclampsia.mp. or "maternal hypertension".mp. or "pregnancy*induced hypertension".mp. or "gestational hypertension".mp. | 571 |
| 14 | ("psychological distress" or anxiety or depression or "mental health" or resilience).mp. | 703259 |
| 15 | 11 and 12 | 120 |
| 16 | 12 and 15 | 66 |
| 17 | limit 14 to human | 62 |
| 18 | limit 17 to female | 53 |
| 19 | 18 not (meta-analysis or "systematic review" or conference abstract or "conference review" or editorial or commentary or correspondence or letter or note or review or book review or news).mp. | 44 |
| 20 | limit 19 to (clinical trial or randomized controlled trial or controlled clinical trial or multicenter study) [Limit not valid in APA PsycInfo; records were retained] | 44 |

[mp = title, abstract, heading word, drug trade name, original title, device manufacturer, drug manufacturer, device trade name, keyword, floating subheading word, candidate term word]

**CINHAL 1985 to 2021 August 22**

| # | **Searches** | **Results** |
| --- | --- | --- |
| 1 | (MH "cognitive behavioural therapy+") or "cognitive behavioural therapy" | 2,631 |
| 2 | (MH "psychotherapy+") or psychotherapy or therap*.mp. | 1,897,303 |
| 3 | psychological | 273,031 |
| 4 | behavio*ral | 120,691 |
| 5 | psych*social | 562,728 |
| 6 | 3 OR 4 OR 5 | 782,394 |
| 7 | (MH "mindfulness+") or mindfulness | 9,010 |
| 8 | (MH "breathing exercise+") or "breathing exercise" or "breathing therapy" | 194 |
| 9 | (MH "acceptance and commitment therapy+") or "acceptance and commitment therapy" | 1,004 |
| 10 | (MH "meditation+") or meditation | 7,192 |
| 11 | "dialectical behavio*r therapy" | 578 |
| 12 | "metacognitive therapy" | 99 |
| 13 | "schema therapy" | 143 |
| 14 | 11 or 12 or 13 | 808 |
| 15 | (MH "exercise+") or exercise or "physical activity" or "mind*body" or yoga | 285,013 |
| 16 | 1 or 2 or 6 or 7 or 8 or 9 or 10 or 14 or 15 | 2,588,194 |
| 17 | (MH "preeclampsia+") or "pre*eclampsia" or "maternal hypertension" or "pregnancy*induced hypertension" or "gestational hypertension" | 9,846 |
| 18 | "psychological distress" | 13,265 |
| 19 | "anxiety" | 107,041 |
| 20 | (MM "Depression, Postpartum") OR "depression" | 181,685 |
| 21 | (MM "Mental Health") OR "mental health" | 157,880 |
| 22 | "resilience" | 13,858 |
| 23 | 18 OR 19 OR 20 OR 21 OR 22 | 381,046 |
| 24 | 17 and 23 | 188 |
| 25 | 16 and 24 | 113 |
| 18 | limit 17 to Peer Reviewed; Research Article; Human;  Publication Type: Clinical Trial, Journal Article, Proceedings, Randomized Controlled Trial, Research  Sex: Female | 76 |

**Search modes** - Find all my search terms

**Expanders** - Apply equivalent subjects

**MIDIRS: Maternity and Infant Care 22 August 2021**

| # | **Searches** | **Results** |
| --- | --- | --- |
| 1 | ("cognitive behavioural therapy" or CBT).mp. | 100 |
| 2 | psychotherapy.mp. or therap*.mp. | 22233 |
| 3 | (psychological or behavio*ral or psych*social).mp. | 13634 |
| 4 | Mindfulness.mp. | 147 |
| 5 | "breathing exercise" or "breathing therapy" | 0 |
| 6 | ("acceptance and commitment therapy" or ACT).mp. | 2213 |
| 7 | Meditation.mp. | 80 |
| 8 | ("dialectical behavio*r therapy" or "metacognitive therapy" or "schema therapy").mp. | 1 |
|  | "social support".mp. | 5929 |
|  | counsel*ing.mp. or psycho*education.mp. | 7951 |
| 9 | (exercise or "physical activity" or mind*body or yoga).mp. | 3417 |
| 9 | 1 or 2 or 3 or 4 or 5 or 6 or 7 or 8 or 9 or 10 or 11 | 16038 |
| 10 | (preeclampsia or pre*eclampsia or "maternal hypertension" or "pregnancy*induced hypertension" or "gestational hypertension").mp. | 7112 |
|  | ("psychological distress" or anxiety or depression or "mental health" or resilience) | 14207 |
|  | 13 and 14 |  |
| 11 | 12 and 15 | 129 |
| 12 | 16 not (meta-analysis or "systematic review" or conference abstract or "conference review" or editorial or commentary or correspondence or letter or note or review or book review or news).mp. | 34 |

[mp=abstract, heading word, title]

**Web of Science – Science Citation Index Expanded (SCI-EXPANDED), Social Sciences Citation Index (SSCI), Emerging Sources Citation Index (ESCI): 1900 to 2021-07-18**

| # | **Searches** | **Results** |
| --- | --- | --- |
| 1 | ("cognitive behavioural therapy" or CBT) | 19,748 |
| 2 | psychotherapy OR therap* | 4,142,892 |
| 3 | (psychological or behavio*ral or psych*social) | 1,201,649 |
| 4 | mindfulness | 19,424 |
| 5 | "breathing exercise" or "breathing therapy" | 462 |
| 6 | ("acceptance and commitment therapy" or ACT) | 1,172,959 |
| 7 | meditation | 12,090 |
| 8 | "dialectical behavio*r therapy" or "metacognitive therapy" or "schema therapy" | 2,770 |
| 9 | "social support" | 77,536 |
| 10 | counsel*ing or "psycho*education" | 172,213 |
| 11 | (exercise or "physical activit*" or "mind*body" or yoga) | 716,171 |
| 12 | 1 or 2 or 3 or 4 or 5 or 6 or 7 or 8 or 9 or 10 or 11 | 6,888,582 |
| 10 | (preeclampsia or pre*eclampsia or "maternal hypertension" or "pregnancy*induced hypertension" or "gestational hypertension") | 36,687 |
| 11 | ("psychological distress" or anxiety or depression or "mental health" or resilience) | 1,151,841 |
| 12 | 10 and 11 | 542 |
| 13 | 9 and 12 | 219 |
| 14 | 13 NOT Document Types: Letters or Editorial Materials or Meeting Abstracts or Review Articles | 167 |

**Search modes** = All fields

**Expanders:** Apply equivalent subjects

**APA PsycArticles 1894 to 2021-08-18**

| # | **Searches** | **Results** |
| --- | --- | --- |
| 1 | ("cognitive behavioural therapy" or CBT) | 4,578 |
| 2 | psychotherapy | 35,331 |
| 3 | (psychological or behavio*ral or psych*social) | 216,827 |
| 4 | mindfulness | 4,336 |
| 5 | "breathing exercise" or "breathing therapy" | 89 |
| 6 | ("acceptance and commitment therapy" or ACT) | 49,524 |
| 7 | meditation | 3,329 |
| 8 | ("dialectical behavio*r therapy" or "metacognitive therapy" or "schema therapy") | 1,515 |
| 9 | "social support" | 19,189 |
| 10 | counsel*ing or "psycho*education" | 41,213 |
| 11 | (exercise or "physical activit*" or "mind*body" or yoga) | 25,552 |
| 12 | 1 or 2 or 3 or 4 or 5 or 6 or 7 or 8 or 9 or 10 or 11 | 220,290 |
| 13 | (preeclampsia or pre*eclampsia or "maternal hypertension" or "pregnancy*induced hypertension" or "gestational hypertension") | 76 |
| 14 | ("psychological distress" or anxiety or depression or "mental health" or resilience) | 94,650 |
| 15 | 10 and 11 | 67 |
| 16 | 12 and 15 | 67 |
| 14 | NOT (Literature Review AND Meta Analysis AND Systematic Review) | 56 |

Limited to peer reviewed literature

**Cochrane Central Register of Controlled Trials Issue 8 of 12, Pregnancy and Childbirth Group**

| # | **Searches** | **Results** |
| --- | --- | --- |
|  | ((("cognitive behavioural therapy" or CBT) OR (psychotherapy OR therap*) OR psychological or behavio*ral or psychosocial OR mindfulness OR "breathing exercise" OR "breathing therapy" OR ("acceptance and commitment therapy" or ACT) OR meditation OR "dialectical behavio*r therapy" or "metacognitive therapy" or "schema therapy" OR "social support" or counsel*ing or "psycho*education" OR exercise or "physical activit*" or "mind*body" or yoga) AND (("psychological distress" or anxiety or *depression or "mental health" or resilience) AND (preeclampsia or pre*eclampsia or "maternal hypertension" or "pregnancy*induced hypertension" or "gestational hypertension"))) | 45 |

**Search modes** – All text (Searched word variations)

**No limit on publication date**
