## Supplementary Table S2 for "Effectiveness of psychosocial interventions for hypertensive disorders in pregnancy: A systematic review and meta-analysis"

### **Appendix S2: Assessment of methodological quality**

**Cochrane Effective Practice and Organisation of Care (****EPOC) Risk of Bias Tool – *EPOC modified tool for assessing risk of bias for randomised trials.***

| **EPOC Quality Assessment Form /Risk of Bias Tool -** Part A | | | |
| --- | --- | --- | --- |
| **Part A** assesses the risk of bias that may be encountered when recruiting participants; allocating to intervention and control groups; inadequate implementation of the intervention; and confounding. Using the guidance provided at the end of this form, select either “high”, “low” or “unclear” for each judgment. When complete, proceed to **Part B** of the quality assessment form | | | |
| **REF ID:** | | | |
| **Domain** | **High risk of bias** | **Low risk of bias** | **Unclear risk of bias** |
| ***1.*** ***Random sequence generation*** | Score “High risk” when a non-random method is used (e.g. performed by date of admission). Non-randomised trials and controlled before-after studies should be scored “High risk”. | Score “Low risk” if a random component in the sequence generation process is described (e.g. Referring to a random number table). | Score “Unclear risk” if not specified in the paper. |
| ***2. Allocation concealment*** | Controlled before-after studies should be scored “High risk”. | Score “Low risk” if the unit of allocation was by institution, team or professional and allocation was performed on all units at the start of the study; or if the unit of allocation was by patient or episode of care and there was some form of centralised randomisation scheme, an on-site computer system or sealed opaque envelopes were used. | Score “Unclear risk” if not specified in the paper.  . |
| ***3. Baseline characteristics similar*** | Score “High risk” if there is no report of characteristics in text or tables or if there are differences between control and intervention providers. Note that in some cases imbalance in patient characteristics may be due to recruitment bias whereby the provider was responsible for recruiting patients into the trial. | Score “Low risk” if baseline characteristics of the study and control providers are reported and similar. | “Unclear risk” if it is not clear in the paper (e.g. characteristics are mentioned in text  but no data were presented). |
| ***4. Knowledge of the allocated interventions adequately prevented during the study ^1,^****^2^* | Score “High risk” if the outcomes were not assessed blindly. | Score “Low risk” if the authors state explicitly that the primary outcome variables were assessed blindly, or the outcomes are objective, e.g. length of hospital stay. Primary outcomes are those variables that correspond to the primary hypothesis or question as defined by the authors. | Score “Unclear risk” if not specified in the paper |
| ***5. Other risks of bias***  ***Bias due to problems not covered elsewhere in the table.*** | Score “High risk” if any important concerns about bias not addressed above. If questions/entries were pre-specified in the study’s protocol, responses should be provided for each question/entry. | Score “Low risk” if there is no evidence of other risk of biases | Score “Unclear risk” if there may be a risk of bias, but there is either insufficient information to assess whether an important risk of bias exists; or insufficient rationale or evidence that an identified problem will introduce bias. |

| **EPOC Quality Assessment Form /Risk of Bias Tool -** Part B | | | |
| --- | --- | --- | --- |
| Part B of this form will assess the Risk of bias for the domains for each group of outcomes. Please indicate the specific outcome and complete the assessment for each. | | | |
| **OUTCOME(S):** | | | |
| **Domain** | **High risk of bias** | **Low risk of bias** | **Unclear risk of bias** |
| ***6. Protection against contamination*** | Score “High risk” if it is likely that the control group received the intervention (e.g. if patients rather than professionals were randomised or there was evidence of interaction between the two groups) | Score “Low risk” if allocation was by community, institution, or practice, and it is unlikely that the control group received the intervention. | “Unclear risk” if professionals were allocated within a clinic or practice and it is possible that communication between intervention and control professionals could have occurred (e.g. physicians within practices were allocated to intervention or control) |
| ***7. Selective outcome reporting*** | Score “High risk” if some important outcomes are subsequently omitted from the results. | Score “Low risk” if there is no evidence that outcomes were selectively reported (e.g. all relevant outcomes in the methods section are reported in the results section). | Score “Unclear risk” if not specified in the paper. For further information see Chapter 13 of the Cochrane handbook: Assessing risk of bias due to missing results in a synthesis |
| ***8. Baseline outcome measurements similar^1,3^*** | Score “High risk” if important differences were present and not adjusted for in analysis. | Score “Low risk” if performance or patient outcomes were measured prior to the intervention, and no important differences were present across study groups. In randomised trials, score “Low risk” if imbalanced but appropriate adjusted analysis was performed (e.g. Analysis of covariance). | If randomised trials have no baseline measure of outcome, score “Unclear risk”. |
| ***9. Incomplete outcome data^1^*** | Score “High risk” if missing  outcome data was likely to bias the results. | Score “Low risk” if missing outcome measures were unlikely to bias the results (e.g. the proportion of missing data was similar in the intervention and control groups or the proportion of missing data was less than the effect size i.e. unlikely to overturn the study result). | Score “Unclear risk” if not specified in the paper (Do not assume 100% follow up unless stated explicitly). |
| ^1^ If some primary outcomes were imbalanced at baseline, assessed blindly or affected by missing data and others were not, each primary outcome can be scored separately.  ^2^ This refers to blinding of participants and personnel and blinding of outcome assessment.  ^3^ If “Unclear risk” or “High risk”, but there is sufficient data in the paper to do an adjusted analysis (e.g. Baseline adjustment analysis or Intention to treat analysis) the criteria should be re scored as “Low risk”. | | | |

Effective Practice and Organisation of Care (EPOC). EPOC resources for review authors: suggested risk of bias criteria for EPOC reviews. EPOC. 2017. http://epoc.cochrane.org/resources/epoc-resources-review-authors.
